# Transformed time series analysis of first-wave COVID-19: universal similarities found in the Group of Twenty (G20) Countries

**DOI:** 10.1101/2020.06.11.20128991

**Authors:** Albert S. Kim

## Abstract

As of April 30, 2020, the number of cumulative confirmed coronavirus disease 2019 (COVID-19) cases exceeded 3 million worldwide and 1 million in the US with an estimated fatality rate of more than 7 percent. Because the patterns of the occurrence of new confirmed cases and deaths over time are complex and seemingly country-specific, estimating the long-term pandemic spread is challenging. I developed a simple transformation algorithm to investigate the characteristics of the case and death time series per nation, and described the universal similarities observed in the transformed time series of 19 nations in the Group of Twenty (G20). To investigate the universal similarities among the cumulative profiles of confirmed cases and deaths of 19 individual nations in the G20, a transformation algorithm of the time series data sets was developed with open-source software programs. The algorithm was used to extract and analyze statistical information from daily updated COVID-19 pandemic data sets from the European Centre for Disease Prevention and Control (ECDC). Two new parameters for each nation were suggested as factors for time-shifting and time-scaling to define reduced time, which was used to quantify the degree of universal similarities among nations. After the cumulative confirmed case and death profiles of a nation were transformed by using reduced time, most of the 19 nations, with few exceptions, had transformed profiles that closely converged to those of Italy after the onset of cases and deaths. The initial profiles of the cumulative confirmed cases per nation universally showed 3–4 week latency periods, during which the total number of cases remained at approximately ten. The latency period of the cumulative number of deaths was approximately half the latency number of cumulative cases, and subsequent uncontrollable increases in human deaths seemed unavoidable because the coronavirus had already widely spread. Immediate governmental actions, including responsive public-health policy-making and enforcement, are observed to be critical to minimize (and possibly stop) further infections and subsequent deaths. In the pandemic spread of infectious viral diseases, such as COVID-19 studied in this work, different nations show dissimilar and seemingly uncorrelated time series profiles of infected cases and deaths. After these statistical phenomena were viewed as identical events occurring at a distinct rate in each country, the reported algorithm of the data transformation using the reduced time revealed a nation-independent, universal profile (especially initial periods of the pandemic spread) from which a nation-specific, predictive estimation could be made and used to assist in immediate public-health policy-making.

Research in context

Evidence before this study
The open data set were obtained from the website of the European Center for Disease Control and Prevention (ECDC). Although the data include the number of new cases and deaths per day per nation, extracting any apparent correlations between unique time-series of nations in different continents is challenging. Nevertheless, cumulative and non-cumulative statistics are, in principle, equivalent, and hence one can be obtained from the other. Because the non-cumulative profiles report instantaneous variations in the pandemic time series, estimation of future trends by extrapolating recent data is often intractable and limited to short-term extrapolations.

Added value of this study
A data transformation method for the cumulative confirmed cases (CCC) and cumulative confirmed deaths (CCD) was developed and used to directly compare the pandemic statuses of multiple nations, especially G20 nations. This model requires data for the nation with the greatest CCC and CCD (especially, during the initial burst of 90–120 days), which, in the case of the COVID-19 pandemic spread, is Italy. Two parameters for time-shifting (*m*) and time-scaling (*β*) are newly introduced and used to define the reduced time *τ*. After the transformation, most nations’ cumulative profiles converge with those of Italy regardless of their geographical locations.

Implications of all the available evidence
The discovery of the universality of the transformed CCC and CCD profiles of multiple countries provides new insight into analyzing pandemic time series, including the current COVID-19 pandemic spread. By shifting and scaling a nation’s pandemic data into the reduced time frame, the nation’s CCC and CCD profiles can be predicted as long as the reference country’s cumulative data are available in the linear time domain. After the extraction of meaningful information from the transformed data, the overall implication is that most nations will reach the same state as Italy’s current state soon, depending on a specific nation’s population and human dynamics.

## Introduction

### A brief history of the first deaths

On December 31, 2019, Chinese health authorities treated a patient cluster of pneumonia caused by the newly recognized coronavirus, i.e., severe acute respiratory syndrome–coronavirus 2 (SARS-CoV-2):^1^ they had closely monitored the cluster from the beginning of December, 2019. The first domestic death in China was reported on January 11, 2020, and afterward the city of Wuhan, with a population of 11·08 million, was locked down. Outside China, the first death was reported by the Philippines on February 2, 2020, followed by France on February 14, Italy on February 22, and the U.S. on February 29. This rapid transmissibility of the coronavirus was estimated by using the instantaneous reproduction number (*R*_*t*_) and confirmed case-fatality risk for four megacities and multiple provinces reporting the highest number of confirmed cases in China.^2^ The aggressive non-pharmaceutical interventions reduced only the first wave of COVID-19 outside of Hubei, and this effort that might have been more successful if foreign importation had been limited to prevent viral reintroduction. Effects of various non-pharmaceutical intervention attempts were reported to be effective in reducing the transmission of COVID-19 (as well as influenza) in Hong Kong, such as border restriction, quarantine and isolation, and social distancing.^3^ Further nation-specific situations of G20 nations can be found elsewhere: Argentina,^4^ Australia,^5,6^ Brazil,^7^ Canada,^8^ China,^9,10,11^ German,^12^ France,^13^ Indonesia,^14^ India,^15^ Italy,^16,17,18^ Japan,^19^ Korea,^20^ Mexico,^21,22^ Russia,^23^ Saudi Arabia,^24^ South Africa,^25,26^ Turkey,^27^ UK,^28^ US,^29^ and multiple European nations.^30^

Because the failure of non-pharmaceutical intervention was ascribed to overseas travel, a global metapopulation disease transmission model was used to project how travel limitations contributed to the mitigation of the global COVID-19 spread.^31^ Within 2 weeks after the first death in China on January 11, the coronavirus appeared to have already been transmitted to other major cities within mainland China.^32^ The reported data suggest that non-pharmaceutical interventions were effective only within cities within China but did not significantly affect the transport of COVID-19 overseas. As of April 30, 2020, the global number of confirmed cases and deaths exceeded 3·3 million and 233 thousand, respectively. Nevertheless, the initial routes of COVID-19 transmission from China to other countries are not well identified, and the correlations among patterns of the occurrence of confirmed cases and deaths in each nation are still ambiguous.

Recent modeling studies used domestic and international CCC profiles, as briefly discussed as follows, but no research was found for statistical characterization of global CCD profiles. CCC of four Asian countries (China, Iran, the Philippines, and Taiwan) from January 21 to April 10, 2020, was investigated using the generalized logistic model (GLM). The results indicated that their CCC profiles follow sigmoidal cures, each with a single turning point,^33^ which seems to be over-simplified. Rapidly increasing CCC numbers in the European Union/European Economic Area and the United Kingdom from January 1 to March 15, 2020, showed similar trends at different stages of the rapid pandemic progress:^34^ No specific mathematical correlations were found. An exponential growth model was proposed to fit critical care admissions to determine likely COVID-19 case numbers (daily new occurrences), critical care admissions, and epidemic growth in the United Kingdom until March 23, 2020.^35^ As the exponential function has the Eigen characteristics, this finding implies that their CCC profile also follows the exponential growth. China’s CCC was modeled using an exponential growth model for the short period from January 20 through 29, 2020, and the strong governmental intervention was modeled using a conditional exponential decay model.^36^ A long-term prediction of China’s CCC was modeled using a method called numbers of the accumulative confirmed patients (NACP). NACP is a linear superposition of multiple sigmoidal functions that fit China’s plateau phase reasonably well. On the other hand, the CCC’s power-law growth until March 27, 2020, was shown to be a promising, descriptive scenario for Brazil, China, France, Germany, Italy, Japan, Spain, Republic of Korea, and the USA in four continents.^37^ Interestingly, China’s short-term CCC was fitted using a fourth-order polynomial of time, defined as the number of days from January 21 to February 23, 2020.^38^ Because a polynomial function usually does not have an asymptotic limit, extending this model for the long-term trend is questionable. Although these studies modeled CCC trends using various mathematical functional approximations at different time and space scales, specific correlations of nations’ pandemic trends were not found.

The objective of this study, therefore, was set to analyze the CCC and CCD of a number of selected countries and investigate any possible universal similarities in the patterns of increases in CCC and CCD over time from December 31, 2019 until April 30, 2020. The observed universal similarities were used to predict a few cases in mid May, 2020. For this purpose, a simple mathematical data transformation model was conceptually developed and implemented by using open-sourced software packages and utilities.

## Methods

### Data collection

All data sets used for this work were downloaded from the ECDC website^39^ as a comma-separated value (csv) format file, named “`download`” without a file extension. The data file contained the daily numbers of confirmed cases and deaths for 206 locations, according to country or territory codes. Open-sourced utilities such as `bash`, ^40^ `sed`, ^41^ and `awk` ^42^ were used as needed to extract the daily CCC and CCD data for individual countries or territories. A total of 206 files were generated, with file names identical to specific countries or territories. Each extracted csv-format file had rows of six selected integer items (i.e., columns): date, day, month, year, cases, and deaths, where the date format was dd/mm/yyyy. In `Octave` (open-sourced software, an alternative clone to MATLAB),^43^ the CCC and CCD were calculated and plotted against the number of days elapsed after December 31, 2019. For each country or territory, eight graphs – new (daily) vs. cumulative, cases vs. death, and linear vs. logarithmic (with base 10) profiles – were plotted against the number of days. The script-generated graphs were automatically saved as image files for visual investigation after the data were plotted. On a desktop computer (Linux OS, Ubuntu 18·04·4 LST (Bionic Beaver), Intel(R) Xeon(R) CPU E5-2697 v4 @ 2·30GHz, 64 GB RAM), this task required only 3–4 minutes for all 206 countries and territories. Close visual inspection of these graphs per nation provided an initial understanding of each nation’s CCC and CCD time series in 2020, where January 1 was set as day 1. Although the basic data extraction and mining were completed for all nations by using the daily updated data file, analysis of all 206 countries or territories was challenging. To potentially include nations representing all continents (excluding Antarctica), 19 individual nations were selected in G20 for the current analysis. The European Union (EU) was excluded because it contains multiple member countries, and ECDC reports COVID-19 data only for individual countries and territories. Extension of the current study to all 206 nations or territories, and even to local areas such cities and communities is straightforward as long as data sets are available. The original csv-format data file from ECDC had, as mentioned above, the time information in each row, including date (dd/mm/yyyy) representation followed by day, month, and year. An open-sourced spreadsheet program called LibreOffice^44^ Calc (an alternative to Microsoft Excel) was used to incorporate the 19 nations’ data with one nation per sheet, and the data analyses were performed with the available functions embedded in Calc. The date-information in “dd/mm/yyyy” format was converted into “yyyy-mm-dd” format to calculate the number of days between two specific dates by using the “DAYS” function, embedded in Calc software. This time conversion from lexical date format to number of days (as a countable integer) was an important basic step for further data investigation and analysis.

### Transformation of time series

#### Target data for investigation

In this work, the cumulative data of cases and deaths (i.e., CCC and CCD) were primarily used instead of data of new daily occurrences with respect to the number of days after December 31, 2019. In principle, using the cumulative information in statistical analysis is equivalent to using a cumulative density function as an integral of a probability density function with respect to control variables. The fundamental advantages of using the cumulative data are as follows. First, the daily data often fluctuate too much to capture specific variations and trends in target variables, so that statistically meaningful characteristics not only are subject to the data observer’s viewpoint but also are often challenging to extract. Second, the cumulative profiles never decrease, so that either rapid/gradual or local/global variations can be systematically captured by using semi-log plots, i.e., common-logarithmic CCC and CCD on the *y −* axis vs. the linear day number on the *x −* axis. Third, the ever-increasing trend in logarithmic cumulative data is often considerably smoother than the original daily time series. Therefore, variation trends can be captured without significant statistical noise. In addition, with a fixed time interval, i.e., 1 day in the current case, the original time series data can be easily retrieved by calculating the difference of the cumulative data between two consecutive days.

#### Formulas for CCC and CCD

For a consistent analysis, we define new variables, such as *C* (*n*) and *D* (*n*) of day *n*, indicating the CCC and CCD, respectively, where *n* increases from *n* = 1 of January 1, 2020 to the day for which the latest data are available. The time series analysis in this study primarily used the cumulative data from January 1 to April 30, 2020, and the developed algorithm was tested using the data from May 1 to 15, 2020. Because the time series of cases and deaths are updated daily, a unit time interval is set as 1 day, i.e., *δn* = 1. Then, CCC and CCD are represented as functions of day number *n*:

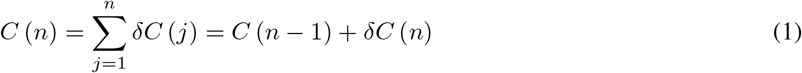

and

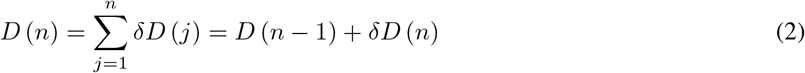

respectively, where *δC* (*n*) and *δD* (*n*) are the increased numbers of cases and deaths, respectively, on day *n*. Some nations do not have complete data sets: the missing days either are in early January, when no pandemic effects were found, or are intermittent 2-3 days after the pandemic report was started. Because cumulative data are processed and analyzed in this study, the missing days are treated as days with no new occurrences, which is to avoid arbitrarily altering the statistical results by interpolation processes.

#### Analogy to statistical physics (thermodynamics)

Data similarity was found unexpectedly between the infection and death time series available online and the phase transition patterns of matter in thermodynamics. In statistical physics, the Clausius-Clapeyron equation describes a discontinuous transition between two phases of single-constituent matter. Various materials have their unique material properties, often represented by using material constants, for example, evaporation enthalpy called latent heat at a specific temperature. Although many organic or inorganic solutions have various trends in their phase-diagrams, e.g., pressure vs. temperature (*P − T*) curves, the overall pattern of how *P* increases with temperature *T* is universal in physics. Along the liquid-gas equilibrium, *P* and *T* are related as

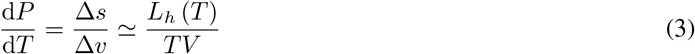

where Δ*s* = *L*_*h*_ (*T*) */T* is the molar entropy change from liquid to gas phases, *L*_*h*_ [Joule/mol] is the molar latent heat as a function of absolute temperature *T* [K], and Δ*v* [liter/mol] is the molar volume difference per molecule between the two phases, which is is often approximated as the gas-phase molar volume. Recent analysis of the Clausius-Clapeyron equation for water has been described elsewhere in detail.^45^ Theoretical ideas for the current work stem from fundamental thermodynamics and statistical physics that the Clausius-Clapeyron equation provides a universal functional form that covers a number of materials undergoing transitions between two or more phases.

#### Reduced times for CCC and CCD per nation

Among the 19 nations belonging to G20, the cases and deaths reported in Italy were selected as principal data. Furthermore, we hypothesized that the CCC and CCD profiles of other nations would have certain degrees of similarity to those of Italy. Because minimizing deaths is a more immediate task than reducing the number of confirmed cases, we investigated the CCD data first. To directly compare the CCD time series of Italy (IT) and that of nation *X*, a characteristic day is defined as the first day when Italy’s CCD exceeded a certain threshold number selected here as 10, denoted *ν*_*d*_ = 10. The day number of the date when Italy’s total deaths exceeded the threshold is denoted 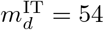 (indicating February 26, 2020). Then, the *x−*axis of Italy’s CCD vs. time graph moves from *n* to 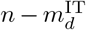 which is equivalent to moving Italy’s profile as many as 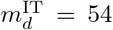 days to the left on the time axis. In general, reduced time *τ*_*d*_ (for the total deaths), specifically for Italy, is defined as 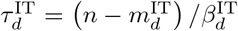 where 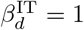 is set by definition because Italy’s CCD data form an international baseline of CCD and therefore do not need to be scaled. A general definition of the reduced time CCD is

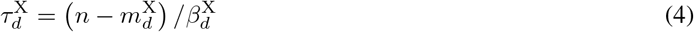

for nation *X*, which will be replaced by the two-character abbreviations of nations investigated.

In this case, 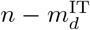 indicates the number of days after the sudden increase in Italy’s CCD. The universal value *ν*_*d*_ = 10 is determined, as it is frequently done for order-parameter estimations in statistical physics, because Italy’s CCD drastically increases after it exceeds 10. A negative value of 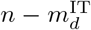 indicates the number of days before the explosive CCD onset. For nation *X*, the reduced time 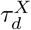 can be obtained by identifying the nation’s day number of CCD onset 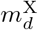, interpreted as a time-shifting parameter, and calculating 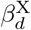, defined as a time-scaling parameter. The physical meaning of 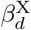 is explained as if 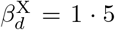; then the CCD rate of nation *X* is 1·5 times slower than that of Italy. In this study, it was found that most nations (except a few outliers) have *β* values greater than 1·0. After 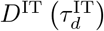 is plotted with respect to 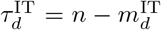 with 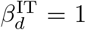, nation *X*’s CCD profile, 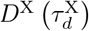, is plotted on the same graph with respect to the nation’s reduced time 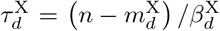 by using an initial guess of 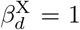. Italy’s variation in CCD has the stiffest slope among all countries after its first COVID-19-related death was reported on February 22, following France’s first reported death on February 14. In this regard, the CCD profiles of other countries, especially the five European nations of Germany (DE), France (FR), Russia (RU), Turkey (TR), and the United Kingdom (UK), are on the right-hand side in the time axis of *n* to Italy’s CCD with lower CCD profiles. While 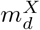 moves the CCD profile of nation *X* to the left to match its onset to that of Italy, the 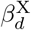 value proportionally shortens (or lengthens) the CCD profile of nation *X* on the shifted axis of 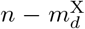. The CCD profiles of Italy and nation *X* are numerically integrated with *n* and 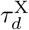, respectively, from 0 to 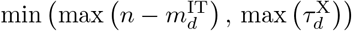. Values of the integrals are the areas under the CCC curves for Italy and nation *X*, whose absolute difference is minimized by iterative adjustment of 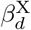 in Calc software. That is, the optimal value of 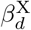 maximizes the overlapping degree of Italy’s linear CCD (without use of the time-scaling parameter 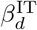) and nation *X*’s transformed CCD profiles. Because 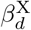 is determined by comparing two finite integrals, 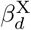 is independent of the pre-selected values of 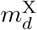. Moreover, CCC profiles are analyzed with the same method for CCD profiles, i.e., for nation *X*, identification of the CCC onset day, calculation of time-shifting parameter 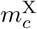, and determination of the time-scaling parameter 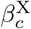. The reduced time for nation *X*’s CCC is similarly defined as

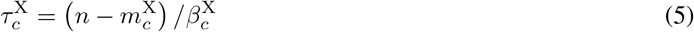

where *m*_*c*_ and *β*_*c*_ are different for each nation. The threshold of CCC is preset as *ν*_*c*_ = 100 using the same criteria to preset *ν*_*d*_. Specific values of *τ* and *β* for CCD and CCC are listed in Table 1 for all G20 nations excluding the EU (denoted G19) nations.

**Table 1:** **Parameters of** the transformation model for G19 nations. In the present study, Turkey is included as a country on the European continent. Although the majority of Turkish land belongs to Asia, Turkey is located closer to Italy than other Asian countries and Russia. Here, 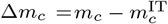 and 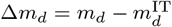 are the time-shifting parameter of a nation, relative to those of Italy.

|  | Nation |  |  | Cases |  |  | Deaths |  |  |
| --- | --- | --- | --- | --- | --- | --- | --- | --- | --- |
| | Abbrev. | Name | Continent | $m_c$ | $\Delta m_c$ | $\beta_c$ | $m_d$ | $\Delta m_d$ | $\beta_d$ |
| 1 | AR | Argentina | South America | 74 | 20 | 2.945 | 85 | 27 | 3.524 |
| 2 | AU | Australia | Australia/Oceania | 69 | 15 | 1.559 | 85 | 27 | 5.467 |
| 3 | BR | Brazil | South America | 70 | 16 | 1.457 | 79 | 21 | 1.538 |
| 4 | CA | Canada | North America | 69 | 15 | 1.626 | 80 | 22 | 1.723 |
| 5 | CN | China | Asia | 17 | -37 | 0.829 | 19 | -39 | 1.396 |
| 6 | DE | Germany | Europe | 59 | 5 | 1.025 | 72 | 14 | 1.565 |
| 7 | FR | France | Europe | 59 | 5 | 1.233 | 68 | 10 | 0.898 |
| 8 | ID | Indonesia | Asia | 72 | 18 | 1.999 | 69 | 11 | 2.256 |
| 9 | IN | India | Asia | 74 | 20 | 1.600 | 83 | 25 | 2.396 |
| 10 | IT | Italy | Europe | 54 | 0 | 1.000 | 58 | 0 | 1.000 |
| 11 | JP | Japan | Asia | 47 | -7 | 3.224 | 70 | 12 | 4.207 |
| 12 | KR | South Korea | Asia | 50 | -4 | 0.914 | 56 | -2 | 2.907 |
| 13 | MX | Mexico | North America | 72 | 18 | 2.152 | 83 | 25 | 1.612 |
| 14 | RU | Russia | Europe | 75 | 21 | 1.353 | 89 | 31 | 2.088 |
| 15 | SA | Saudi Arabia | Asia | 69 | 15 | 2.365 | 88 | 30 | 2.824 |
| 16 | TR | Turkey | Europe (Asia) | 75 | 21 | 0.997 | 76 | 18 | 1.915 |
| 17 | UK | United Kingdom | Europe | 65 | 11 | 1.021 | 73 | 15 | 1.003 |
| 18 | US | United States | North America | 62 | 8 | 1.332 | 67 | 9 | 1.221 |
| 19 | ZA | South Africa | South Africa | 74 | 20 | 1.600 | 83 | 25 | 1.840 |

## Results

### Statistical analysis: Overview

This work was motivated by interesting similarities among the CCD data for six nations in the continent of Europe included in G19: DE, FR, IT, RU, TR, and the UK. These six nations are denoted E6 throughout the manuscript, and E5 denotes all those nations except Italy, i.e., the nation with the highest number of CCC and CCD at most times to date. Figure 1 shows the pandemic time series of the E6 nations: CCC profiles on (a) linear and (b) logarithmic scales and CCD series on (c) linear and (d) logarithmic scales. Notably, the *y*−axis maxima of (a) and (b) for CCC are much higher (approximately a tenfold or more) than those of (c) and (d) for CCD. The current fatality rate of COVID-19 is estimated to be on the order of *O* (10^*−*2^), i.e., a few percent. Herein, CCC and CCD data are plotted on the *y*−axes of the linear and logarithmic (base 10) scales along (linear) *x* axes of time, i.e., the number of days after December 31, 2019, which are denoted in linear and logarithmic plots, respectively. In general, two data lines in a linear plot can be compared by observing the apparent dominance of one line over the other in terms of magnitude. The logarithmic plots allow for comparison of two data lines at various orders of magnitude. For example, the onset-time and latency-period of each nation are better visualized in the logarithmic plots, whereas their one-to-one comparison is more straightforward in the linear plots.

**Figure 1:**
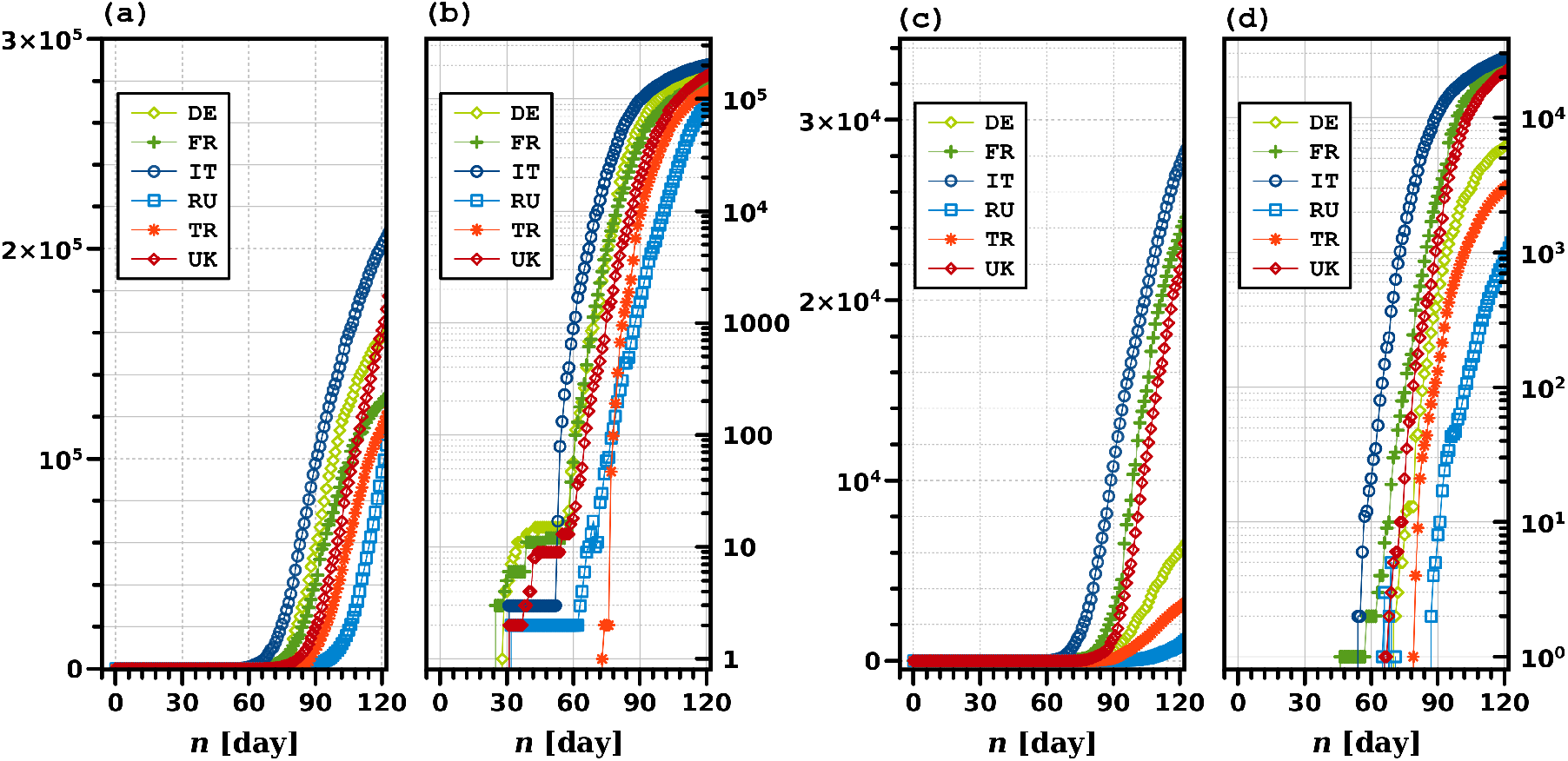
Time series profiles of the six nations (E6) on the European continent: cumulative confirmed cases on (a) linear and (b) logarithmic scales, and cumulative confirmed deaths on (c) linear and (d) logarithmic scales, by April 30, 2020. The day *n* indicates the number of days from December 31, 2019, so that day 1 indicates January 1, 2020.

Several unique aspects identified by simple visual investigation in figure 1 became the major motivation for this study. First, the linear CCC profiles, shown in figure 1(a) appear to follow an ordered previously unknown pattern. Except for the CCC case of the UK, no intersections between two nations are seen. This trend is exceptionless in the CCD profiles shown in figure 1(c), thus strongly implying that, if the ordered pattern continues for all E6 nations, none of the E5 nations will have more severe situations than Italy, which has the highest CCC and CCD numbers. This argument was found to be reasonably valid until mid-April, 2020 (i.e., day 100 or later). Individual nations’ immediate enforcement of public health policies may be able to alter increasing CCC rates, but non-pharmaceutical interventions are known to be inefficient after COVID-19 spread become prevalent. Second, figure 1(b) shows a sudden onset followed by a latency period of approximately 1 month. During this latency period, the CCC remains more or less 10, specifically, between 2 and 20. One exception is Turkey, showing only a few days of CCC latency: Turkey’s CCC also intersects with that of Russia before it exceeds the threshold around 100. Determination of the onset time and latency period of a nation is more straightforward by visually analyzing the cumulative data rather than daily profiles of new occurrences. Third, figures 1(c) and (d) show the E6 nations’ CCD plots on linear and logarithmic scales, respectively. As discussed above, no nations show any noticeable intersections with any other nations within day 120. On the epidemic timeline, this ordered pattern may be limited to neighboring nations during an intermediate-term spread on the order of months. Notably, the CCD profiles are similar between Italy and Russia for the highest and lowest values within the E6: Russia’s CCD can be fitted by shifting Italy’s CCD to the right on the time axis and shrinking the magnitude by multiplying by a constant or a predictable weighting function, or vice versa. In the following sections, Italy’s profiles are kept fixed for consistency while other nations’ profiles are transformed by using a *reduced time* concept. Fourth, although figures 1(b) and (d) appear similar, there are noticeable differences in profile overlap and latency period. As shown in figure 1(c) and (d), the CCD profiles of the E6 nations do not show apparent overlaps. More importantly, the latency periods of CCD profiles are often too short to recognize, and the subsequent death bursts are very abrupt, showing stiff slopes. The onset profiles of the E6’s CCD suggest that after a nation identifies the first death related to and/or caused by the coronavirus, the death burst will inevitably occur within no more than a week. Updated plots of E6’s CCC and CCD profiles until May 15, 2020, are included as figure A.1 in Appendix.

### European nations (E6)

The pandemic profiles of the E5 nations are investigated by calculation of each nation’s *m* and *β* values for CCC and CCD, which are compared with those of Italy as baseline values. Figures 2 (a) and (b) show the apparent similarities between the E5’s transformed profiles and Italy’s linear profiles of CCC and CCD, respectively. This universality of the pandemic time series data is present in both confirmed cases and deaths, because an infection is a necessary condition for death unless complications of the COVID-19 pandemic develop unexpectedly. Since the key process in the current study is transforming the cumulative profiles of nation *X* onto those of the reference country (Italy), the threshold values for CCC and CCD must be preset to calculate *m* and *β* values. These threshold values are subject to intuitive data observation, allowing qualitative human perceptions to be input into numerical calculations for more meaningful data interpretations.

**Figure 2:**
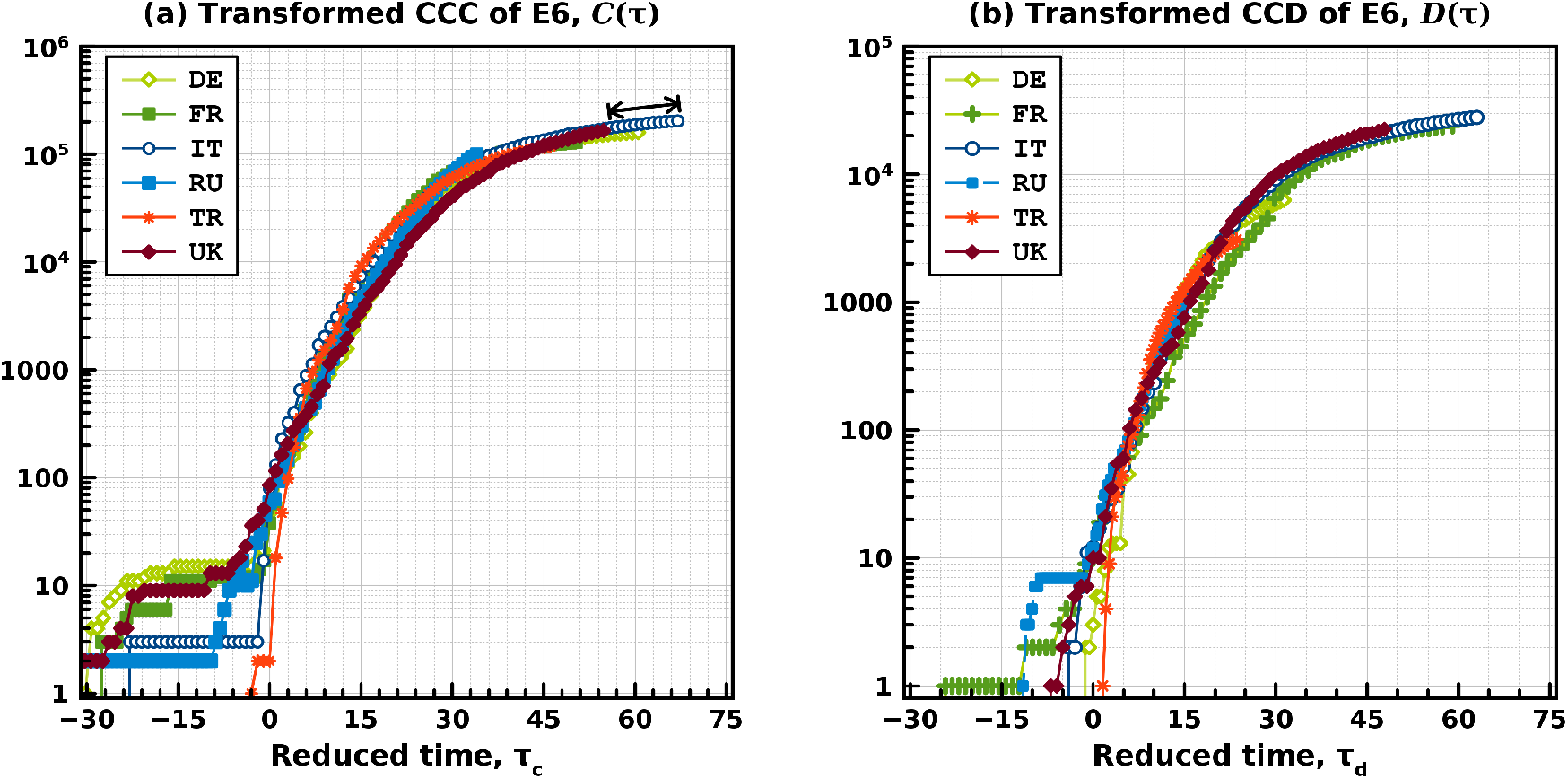
Transformed (a) CCC and (b) CCD profiles of the six nations (E6) in the European continent, by April 30, 2020. The raw data of CCC and CCD are identical to those used on Fig. 1. The reduced time *τ*_*c*_ and *τ*_*d*_ are calculated with Eqs. (4) and (5), respectively, with specific parameter values of *m* and *β* calculated in this study. (See Table 1 for details.) The double arrow toward the end-point of Italy’s profile in (a) indicates the estimated time distance for the U.K. to reach the same CCC level as that of Italy.

### CCC of the E6

Figure 2(a) shows that not all E6 nations have a similar onset trend after a certain latency period, but most appear to follow Italy’s profile after their CCC numbers exceed approximately no more than 100, denoted here as the default CCC threshold *ν*_*c*_ = 100. During the transformation, the actual threshold value of a nation’s CCC is flexibly and manually adjusted to identify the best profile-matching values of *m*_*c*_ and *β*_*c*_ per nation. In general, the best matching is obtained by concurrently searching *ν*_*c*_, *m*_*c*_ and *β*_*c*_ by using the “`Solver`” function in OpenOffice Calc. The target matching zone of the reduced time is where Italy’s CCC varies from 100 to 10,000. Germany, France, and the UK clearly have longer latency periods of approximately 3-4 weeks, and three other nations have latency periods of less than 1 week. Italy’s latency period appears as almost four weeks with a small CCC number of 3. Much in-depth research is required to understand the heterogeneous onset trends in neighboring nations, but the similarity of their transformed CCC profiles after *τ*_*c*_ *>* 0 is, to the best of my knowledge, a unique finding in this work. All E5 nations’ CCC profiles show a tendency to stop somewhere in the middle of Italy’s path. For example, CCC profiles of the UK and Italy can be analyzed by transforming with the parameters of *m*_*c*_ and *β*_*c*_, respectively: 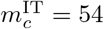 and 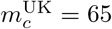 indicates that the UK’s CCC onset is 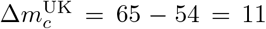 days after that of Italy, as listed in Table 1. This onset time difference is graphically represented as the distance between the highest CCC values of the UK and Italy. In addition, 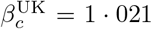 indicates that the propagation speed of the confirmed cases in the UK is 1·021 times (or 2·1 percent) slower than that of Italy, so that the UK and Italy have the same rate of increase in CCC over time. The difference in the reduced time between the latest CCC numbers between Italy and the UK is 67-58·83=12·17 days in figure 2(a), and if these values are multiplied by 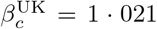, then 12·45 days is obtained, which is the time distance in real days for the UK to reach the highest level seen in Italy. That is, the UK’s CCC will increase from 165 thousand (as of April 30) and reach that of current Italy (203,591 as of April 30) by May 12 or 13 (i.e., 12 or 13 days after April 30). The UK’s CCC on May 12 and 13 are 223,060 and 226,463, respectively, which shows the predictability of this transformation method within approximately 12.8% of error. This time distance of the UK toward Italy is mathematically defined as

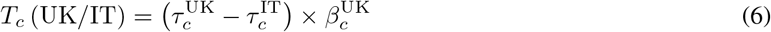

and conceptually visualized by a double arrow at the end of Italy’s CCC profile in 2(a). In addition, Germany and France have CCD onsets 5 days after that of Italy, and France’s CCD is approximately 23.3% slower than that of Italy, but Germany’s CCC propagation is as fast as that of Italy because of 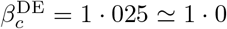. Turkey and Russia both started their CCC onset 21 days after Italy’s: in terms of CCC propagation rates, Turkey 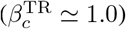 has an equal pace to that of Italy, and Russia 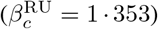 is the slowest among the E6 nations and is close to the US 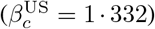. These nation-specific, complex trends of CCC onset and burst are explained using the simple transformation method that can predict the time distance to the future situations using the reduced time concept.

### CCD of the E6

CCD analysis methods are the same as those of CCC, described above. Table 1 indicates that the *β*_*d*_ values of the E5 nations are greater than or (almost) equal to that of Italy, i.e., 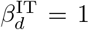 such as 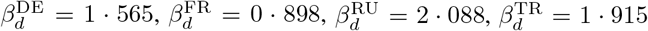, and 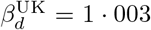. Uniquely, the UK shows a CCC and CCD pace equal to that of Italy, so that 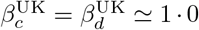 can be reasonably approximated. Germany, Russia, and Turkey, but not France, have higher *β*_*d*_ values than 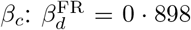, thus indicating that the death rate of France is 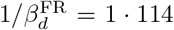 times (or 11·4 percent) faster than that of Italy. Because France is the only nation with *β*_*d*_ *< β*_*c*_, its slower CCC and faster CCD rates than those of Italy may be ascribed to the first death in France occurring 8 days earlier than Italy’s first death on February 22. Similarly to *T*_*c*_ (UK*/*IT), the CCD time distance of the UK, denoted *T*_*d*_ (UK*/*IT) is calculated as

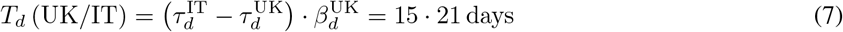

where 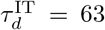 and 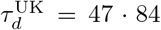 as of April 30. Eq. (7) predicts that from April 30, 15 or 16 days will be required for the UK to reach the same CCD level of Italy as of April 30. On the basis of both *T*_*c*_ (UK*/*IT) ≃ 12 and *T*_*d*_ (UK*/*IT) ≃ 15, in mid-May, the UK is expected to be in a similar situation in terms of CCC and CCD to Italy’s situation as of April 30. On May 15 and 16, the UK’s CCD reached 29,990 and 30,374, which is within 8.5% of error compared to Italy’s CCD of 27,825 on April 30, 2020. This result confirms that a specific nation’s CCC and CCD profiles can be reasonably well predicted using the time distance concept. As already shown in figure 1, the CCD profiles, either linear or scaled, show latency periods too short to be used as precursors for short-term public-health policy enforcement. Although some subtle variations are seen in E6 nations’ CCD time series, E5 nations’ CCD profiles are governed by the same rule and converge to a predetermined destination, i.e., Italy’s current situation, in any future scenario. After April 30, 2020, the UK’s CCC and CCD numbers exceeded those of Italy, as shown in figure A.2(a) and (b), including May data on to figure 2(a) and (b), respectively. Russia appears to start the secondary CCC burst after April 30; however, its CCD record is still one order of magnitude lower than that of Italy because Russia’s first death is 31 days after that of Italy. More one-to-one comparisons between nations and Italy could be used to provide specific information for decision-making but are beyond the scope of this work’s focus on developing and implementing the transformation algorithm with the newly defined time-shifting and time-scaling parameters of *m* and *β*, respectively. Nevertheless, the transformation methods used above for E6 nations are extended to additional nations.

### Nineteen nations in G20 (G19)

We expand the list of nations to test whether the CCC and CCD similarities among E6 nations are also present in more nations on six different continents: Africa, North and South America, Asia, Australia/Oceania, and Europe. In this regard, we select G19 as described above. Because the EU is not an individual nation but a group member of European countries, it is not included in the pandemic analysis in this study. Data used for the G19 analysis implicitly include those of the E6 used above. Figure 3 shows the G19 nations’ linear profiles of CCC and CCD, denoted *C* (*n*) and *D* (*n*), respectively, and their scaled profiles of *C* (*τ*) and *D* (*τ*), respectively.

**Figure 3:**
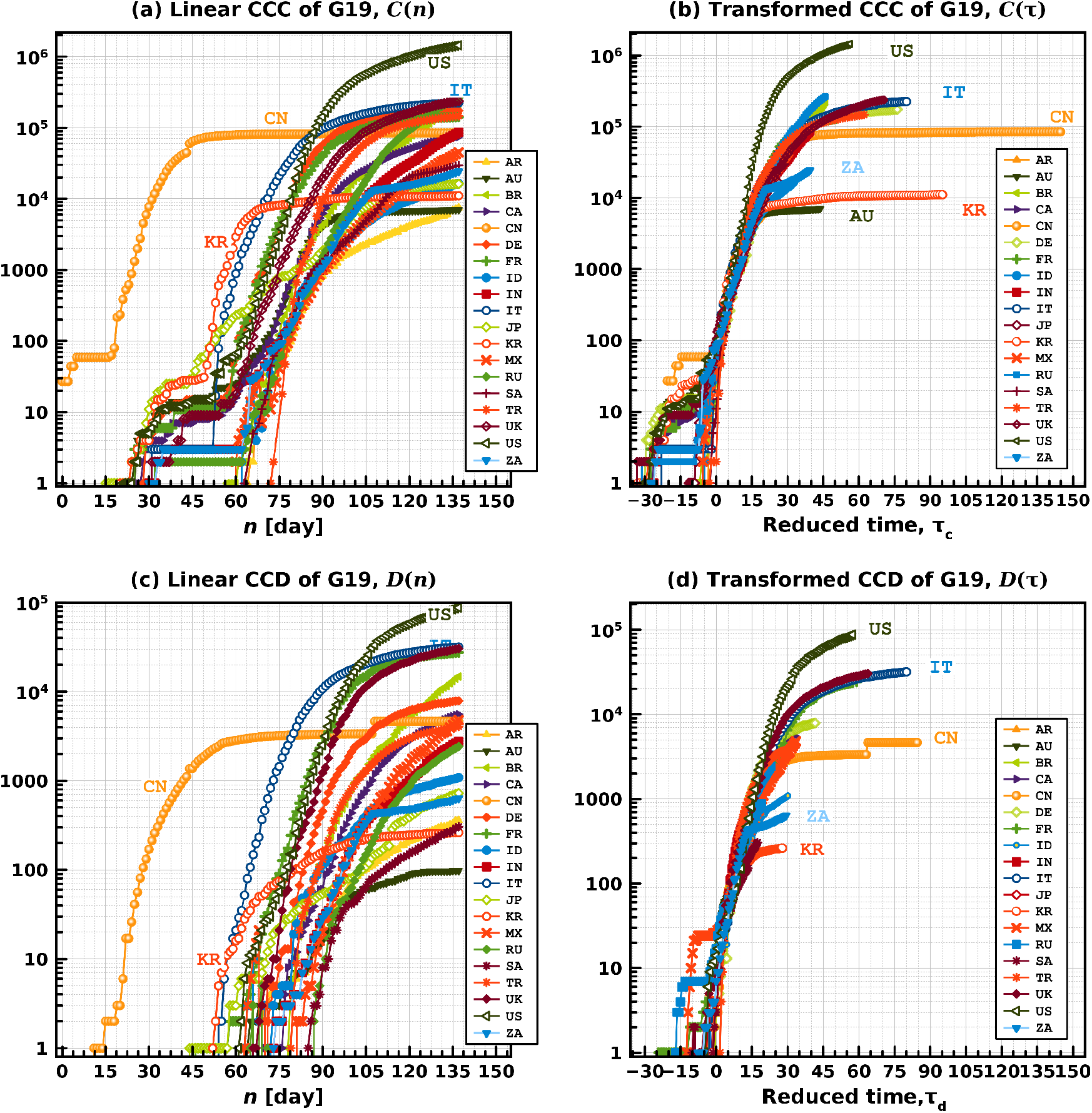
The cumulative confirmed cases plotted on (a) linear and (b) reduced time scales and the cumulative confirmed deaths, plotted on (c) linear and (b) reduced time scales, of all G19 nations (including E6 nations). Two-character abbreviated nation names are used for simplicity and are alphabetically ordered in the legends. To show intersecting profiles in CCC and overlapping profiles in CCD plots, some data lines are overlapped.

### Overview of G19

The overall trend in *C* (*n*) profiles in figure 3(a) is similar to that of *D* (*n*) in figure 4(c) slightly moved to the left in the time axis, because a nation’s CCC profile precedes its CCD profile in time. As previously discussed for the E6 cases, the threshold cut and latency periods of CCC onsets are generally higher and longer than those of CCD onsets, respectively. Most G19 nations show a two-step CCC burst: a first burst from *O* (1) to *O* (10−10^2^), followed by a certain latency period before a second burst from *O* (10−10^2^) to at least *O* (10^4^) or higher. As graphically shown in figures 3(b) and (d), the universality of the cumulative profiles is not limited to E6 nations in the European continent but also appears globally valid for diverse G19 nations in all six continents. The ordered sequences of the E6 profiles shown in figure 1(b) and (d) are absent in figures 3(a) and (c), respectively, so that the ordered patterns of the cumulative profiles must be present, especially in the initial pandemic period of about 90 days, limited to neighboring nations with similar land sizes, populations, and lifestyles in all aspects. The US, China, and South Korea show distinct trends in figures 3(a) and (c); more importantly, both the CCC and CCD profiles of all G19 nations converge to those of Italy for approximately 2-4weeks of reduced times *τ*_*c*_ and *τ*_*d*_ after their onsets, as shown in 3(b) and (d), respectively. Statistically, outlying nations include the US whose highest CCC and CCD exceeded those of Italy on March 27 and April 12, respectively. The earliest onsets of CCC and CCD in China are shown in figures 3(a) and (c), respectively, and their strongly invariant tendencies are shown in 3(a)–(d). Excluding China, South Korea is, to the our knowledge, the only nation that noticeably deviated from the cumulative profiles of Italy ahead of time, and South Africa (ZA) appears to follow South Korea as indicated in figure 3(d). Further monitoring is necessary to estimate the distance of South Africa’s latest cumulative values to Italy’s profiles.

**Figure 4:**
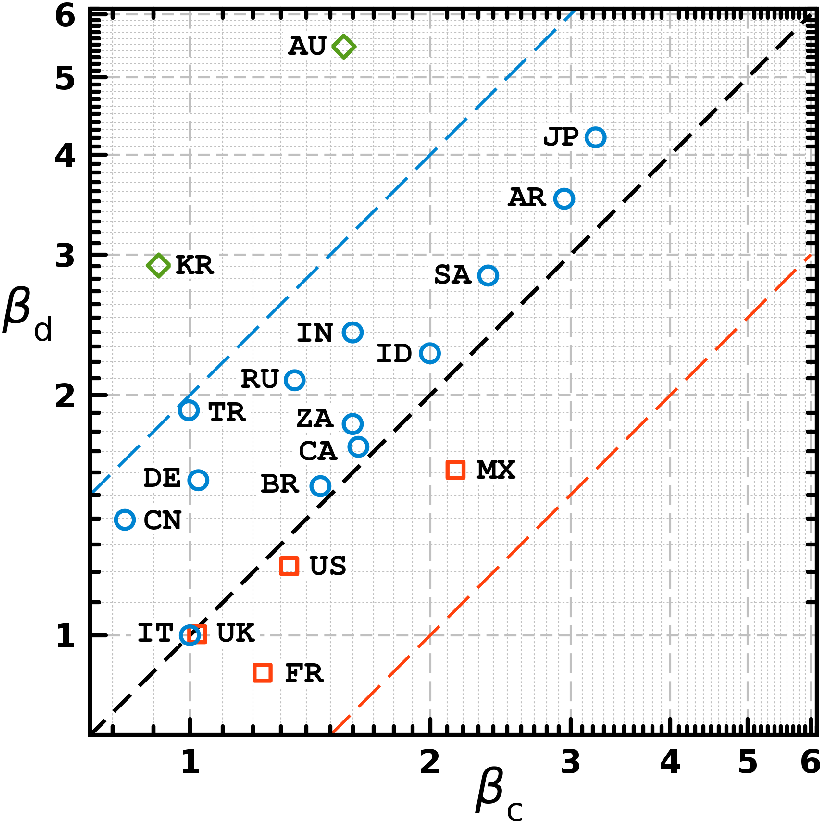
Correlation plots of *β*_*d*_ vs. *β*_*c*_ of G19 nations. The dotted line is the diagonal line of *β*_*d*_ = *β*_*d*_, on which Italy (1,1) is located.

### CCC of G19

In figure 3(b), China shows high similarity in its 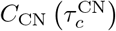 to 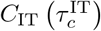 until its CCC approaches 10^5^. The low-level plateau profile of South Korea, 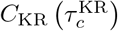, is maintained at approximately 10^4^, i.e., approximately one and two orders of magnitude smaller than those of China and the US, respectively. The front-end of the 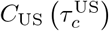 profile in figure 3(b), i.e., in the most recent days, shows a much stiffer slope than those of China, South Korea, and even Italy. It is concerned that 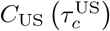 will increase more rapidly than those for any other nations in G19. Except for a few abnormal or outlying CCC profiles, figure 3(b) shows a strong universality among G19 nations in converging to the baseline CCC of Italy, especially during the initial reduced time *τ*_*d*_ of approximately 15 days, despite the large population differences and the different continents.

### CCD of G19

Figure 3(c) confirms the earliest onset of China’s CCD in late January (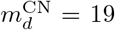 with the default threshold of *ν*_*c*_ = 10) after the first death reported on January 11, 2020. From March onward, the reported CCD data show only small increases, before an abrupt increase due to the reporting of 1,290 new deaths on April 17 (day 108). Figures 3(c) and (d) show that the *D*_KR_ and *D*_IT_ profiles bifurcate and take two distinct routes. *D*_US_ (*n*) and *D*_FR_ (*n*) increase similarly after their onsets until *D*_US_ (*n*) exceeds *D*_FR_ (*n*) and *D*_IT_ (*n*) on March 31 (day 91) and April 12 (day 103), respectively. 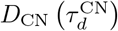 shows a slow increase or even a plateau trend, thus indicating that the new number of deaths is manageable. The universal linearity of all G19 nations shown in 3(d) in the reduced time *τ* from 0 to 30 mathematically implies a simple power law for a nation *i* such as

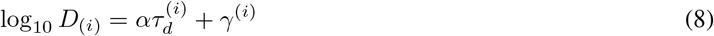

where *α* is the universal slope and *γ*^(*i*)^ is an *y*−intercept. On the basis of Italy’s CCD data from the first 21 days after onset, *α* = 0·121 and *γ*^IT^ = 1·02 are calculated.

### The United States

In figure 3(a) and (c), as of April 30, *C*_US_ (*n* = 121) = 1, 040K is approximately 5·10 times *C*_IT_ (*n*) = 204K, and this ratio is in good agreement with the population ratio (in 2019) of the US (328·2 M) and Italy (60·36 M), i.e., 328 2 60 36 = 5 44. However, the ratio of *D*_US_ (*n*) = 609 · 7K and *D*_IT_ (*n*) = 27 · 8K, i.e., 2·19, is below half the population ratio as of April 30. The other, more rigorous way to estimate the death rate is as follows. The CCD reduced time of the US as of April 30 (day number *n* = 121) is 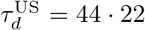, at which 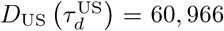. For Italy, 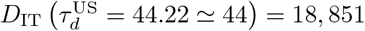. Because 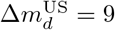, one can find 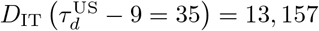. Then, the ratio of

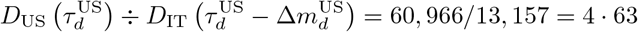

which is closer to *C*_US_ (*n* = 121) */C*_IT_ (*n* = 121) = 5 · 10 and the population ratio of 5·44. Although the ratios calculated above are not a common academic standard but instead are empirical, they provide a quantitative method to estimate the future CCC and CCD of the US, as derived from the baseline values for Italy. The coincidence of the total case and population ratios is limited to the US and further in-depth research is crucial for more fundamental evaluations.

For the US, 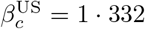 and 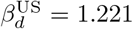 indicate that both the case and death rates are 33·2 percent and 22·1 percent slower than those of Italy, respectively. The CCC and CCD onsets of the US are only 8 and 9 days after those of Italy. However, after the US’s CCD onset, only 38 days (from March 5 to April 12) are required to exceed Italy’s linear CCD profile, *D*_IT_ (*n*), in figure 3(c). The same trend in the US is found in figure 3(d), such that 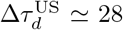 days from 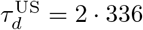 of the onset to 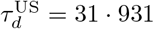 for 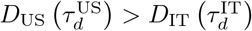. The steady increase in the gap between 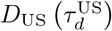 and 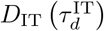 in figure 3(d) implies that 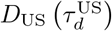 already entered an unprecedented phase that cannot be predicted using the front-end profile of 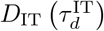. The outlying increases in the US’s CCC profile shown in figure 3(b) should be further analyzed in depth, because the US currently has the largest number of CCD in the world, and, more severely, the greatest potential for further deaths. Closely monitoring the UK, Russia, and Brazil, in addition to the US, indicates that Italy’s role as the reference country may end for this first wave of onset and burst of global cases and deaths. The second wave is not yet predictable using the current knowledge, obtainable from the data released by ECDC.

### Time-scaling parameters of CCC and CCD

Figure 4 shows a scatter plot of *β*_*c*_ vs. *β*_*d*_ for all G19 nations. Italy is, in principle, positioned at (1,1) on the diagonal line. A position above the diagonal line indicates that the nation’s death rate is slower than the infection case rate, i.e., *β*_*d*_ *> β*_*c*_. Most nations are located between the diagonal and upper lines, with the exceptions of Australia and South Korea. In Australia, even if the number of infection cases were to increase as rapidly as those of most nations, such as India, South Africa, and Canada, the death rate is approximately three times slower than the infection rate. South Korea is located at (0.914, 2.907) far above the Italy position (1,1), but below that of Australia. Although South Korea initially had a faster increase in the infection number 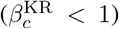 than that of Italy, their death rate significantly decelerates after day 90 in figure 3(c). Three nations, France, Mexico, and the US, are below the diagonal line. Although the positions of these three nations are still near the diagonal line, their death rates are of concern because of *β*_*d*_ *< β*_*c*_. For more meaningful analyses, various domestic conditions in the nations should be considered systematically in addition to the actual number of CCC and CCD; however, such analysis is beyond the scope of this research.

### Role of the funding source

The funder of this study had no role in the study design, data collection, data analysis, data interpretation, or writing of the report. The corresponding author had full access to all the data in the study and had final responsibility for the decision to submit for publication.

## Discussion

The present study used a mathematical transformation to identify universalities among many nations (on multiple continents) lacking apparent similarities in population, land size, and socioeconomic conditions. Two time-related parameters were newly introduced in this study for the CCC CCD: the time-shifting parameter *m* and the time-scaling parameter *β*. These parameters move a nation’s cumulative profile to a new time-origin and allow the nation’s profile to be matched to Italy’s baseline by stretching or shrinking the profile (anchored at the new origin) along the time coordinate (i.e., *x*−axis). The *m* and *β* values were obtained individually for the CCC and CCD of each nation and used to define the reduced time *τ*. By transforming a nation’s data relative to Italy’s baseline CCC and CCD, the short-term estimation of cumulative profiles becomes possible, and the results can be used for broad types of decision-making. Because the large number of individual nations and territories where the COVID-19 pandemic caused severe public health problems, the current study is restricted to the time series of the CCC and CCD of 19 independent nations within G20 (excluding the EU). The primary research idea originated from the sequentially ordered patterns of CCD time series found in six nations on the European continent during the early stage of the pandemic spread, i.e., within 90 days. With the transformation methods of reduced time, both the CCC and CCD profiles of the five European nations converged to those of Italy, which were used as baselines for the rest of the present study. Exceptions observed were China, South Korea, and the US, owing to their noticeable deviations from the CCC and CCD profiles from those of Italy. When the transformation of the cumulative data was extended to all G19 nations, the universality of the profile convergence was found to be valid for as many as 15 nations within G19, excluding the three exceptions above and the reference country, Italy. The common plateau profiles of China’s CCC and CCD, reached in the middle of February 2020, showed early deviation from Italy’s baseline profile. South Korea’s CCC profile appeared to be a down-sized version of China’s, representing only a small number of new recent cases per day. If the CCC profile is assumed to be a good precursor of incoming CCD, South Korea’s CCD profile already have appeared to deviate from that of Italy (see figure 3). On a linear time-scale, the CCC and CCD profiles of the US already exceeded those of Italy in late March and early April, respectively, but the US profiles in the reduced time-scale became the world largest values much earlier than those observed on the linear time-scale. Visual investigation of the transformed CCC and CCD profiles implied that, unlike those of other nations, the US profiles intrinsically did not follow Italy’s profiles but increased much faster over the most recent CCD and CCC values of Italy. A rough but conservative prediction of the US’s CCC is approximately five fold higher than that of Italy, a result similar to the US-to-Italy population density ratio.

More importantly, for future responses to a similar pandemic spread, I emphasize new fundamental insights obtained and findings from the transformation model developed herein. The CCD and CCC profiles of different nations show subtle but distinct characteristics in their onset behaviors. The six nations in Europe had drastic increases in CCD immediately after their onsets. Only France and Russia showed graphically recognizable CCD latency periods. However, five nations in Europe (except Turkey) showed a longer latency period of CCC, close to four weeks. For the six nations in Europe, the threshold values of CCC and CCD were on the order of *O* (10) and *O* 10^2^−10^3^, respectively, which correlated with the fatality rate of COVID-19, close to 7.0% as of April and May, 2020. Similar characteristics of the latency periods and threshold values of CCC and CCD were found for all G19 nations, implicitly including E6 nations. First, in general, the CCC profile of a nation usually has a longer latency period and a higher threshold value than those of CCD of the nation, respectively. Second, there exists a baseline CCC and CCD profiles of a nation whose status are more severe than that of any other nation in the initial pandemic period of at least 90 days; this nation was Italy in the current study. The three outlier nations whose CCD/CCC profiles did not converge to those of Italy were the US,China, andSouth Korea. On the basis of visual investigation of transformed CCC/CCD plots of these nations, the CCC/CCD of the US, which already exceed those of Italy, is expected to increase much faster than those of any other nations, to an unprecedented level. The CCC and especially the CCD of South Korea have almost certainly already stabilized to seemingly constant values, thus indicating that the number of new confirmed cases per day should decrease down to one- or two-digit numbers. Profiles of China has the longest time series with an abrupt daily jump of 1,290 in CCD but no similar variations in its CCC.

In recent engineering disciplines, big data research and applications have become an essential component of future development. In this regard, two representative approaches are *data-driven* and *data-oriented*: in the former, progress is compelled by data, excluding human inputs, whereas the latter is originally intended to optimize software programs against the object-oriented programming of a poor data locality. In decision-making processes during the current global health crisis, a novel paradigm should include the principal advantages of the robust data structures of data-driven approaches, relaxing the excessive dependence on data and ensuring efficient data-utilization in data-oriented approaches for prompt, accurate decision-making. In addition, *data-informed* approaches can be also considered to create proper balances in decision-making by fully utilizing demonstrated knowledge, experience, skills, and predictive ability.

In conclusion, the current study of the mathematical transformation (i) combined three data-utilization approaches by using the pandemic data from ECDC on human infection and death caused by COVID-19 without any mining processes, (ii) developed program scripts by using open-sourced software, and (iii) intuitively applied basic principles of statistical physics to pandemic time series analyses. New outcomes of the current work are the developed transformation model of the reduced time *τ* with two parameters of *m* and *β*, tabulated for the CCC and CCD time series of a total of 19 nations. Specifically, this study suggests that the universality found by using the transformation model can be effectively and efficiently applied in public health policy-making for any nations whose CCC and CCD profiles follow those of the leading nation, i.e., Italy. Because neighboring nations have similar trends in CCC and CCD propagation over time, close international collaborations involving sharing of human and medical resources would effectively decelerate the pandemic spread and risk to human lives.

## Data Availability

The data that support the findings of this study are openly available in https://www.ecdc.europa.eu/en/publications-data/download-todays-data-geographic-distribution-covid-19-cases-worldwide.

## Contributors

ASK designed the study, collected the data, developed scripts of open-sourced software, analyzed and interpreted the data using the scripts, and wrote the manuscript.

## Acknowledgments

This paper was reviewed by Thomas C. Hardy, a professional medical editor, for accuracy and correct language use.

## Nomenclature

CCC: Cumulative confirmed cases
CCD: Cumulative confirmed deaths
DCC: Daily confirmed cases
DCD: Daily confirmed deaths
E5: E6 without Italy
E6: Six nations of Italy, France, Germany, the United Kingdom, Turkey, and Russia, located in the continent of Europe and members of G20.
ECDC: European Center for Disease Control and Prevention
G19: The Group of Twenty excluding the EU
G20: The Group of Twenty

## Appendix

Figures A.1 and A.2 include additional data from May 1 to 15, 2020, onto figures 1 and 2, respectively. During this period, Russia’s CCC rapidly increased over that of Italy, while its CCD is still under Italy’s CCD. The UK’s CCC and CCD have gradually exceeded those of Italy so that the current model with Italy as the reference country might not be as predictable as the present analysis. A population-based pandemic dynamics model is of great necessity.

**Figure A.1:**
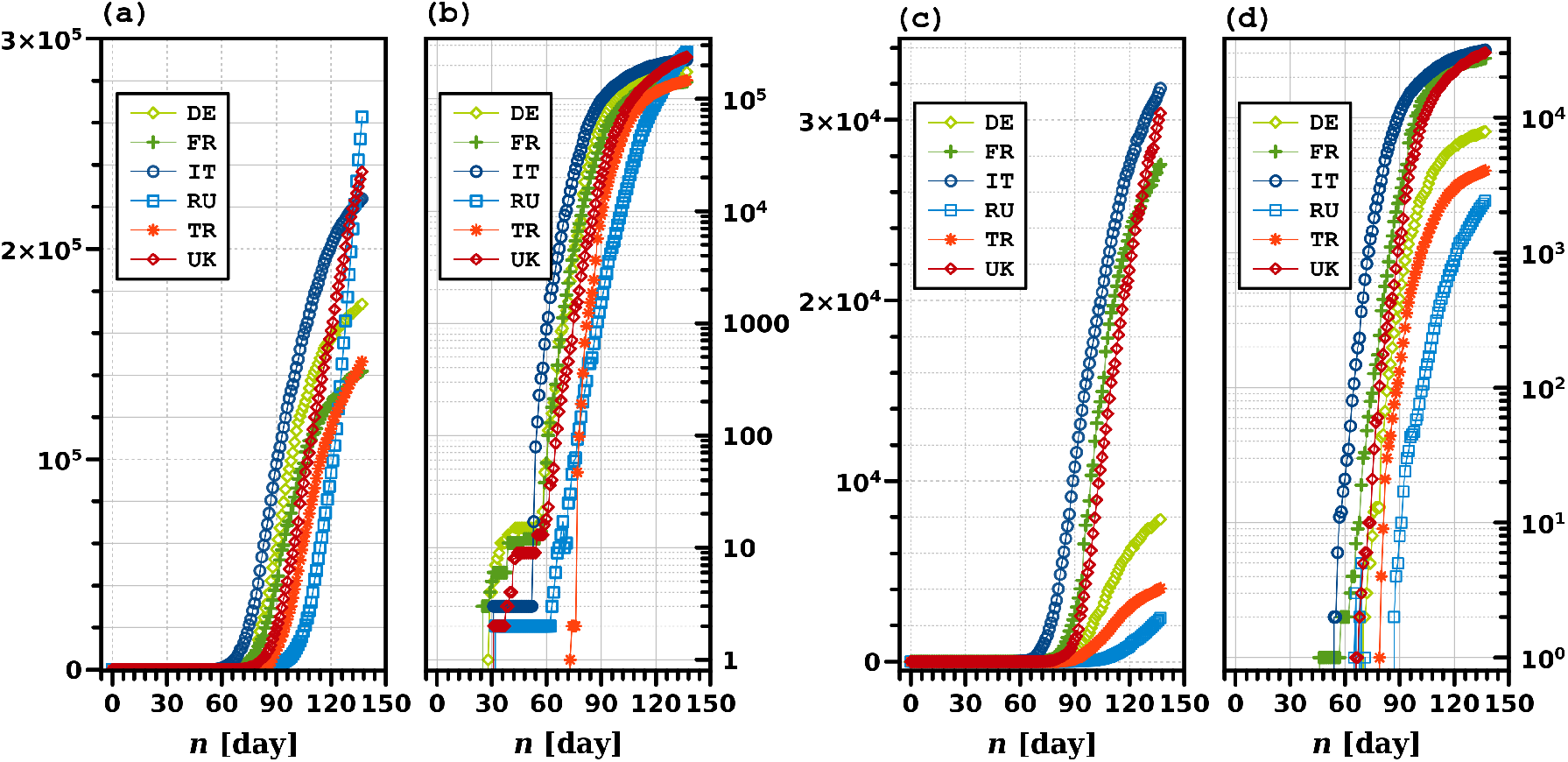
Time series profiles of the six nations (E6) on the European continent: cumulative confirmed cases on (a) linear and (b) logarithmic scales, and cumulative confirmed deaths on (c) linear and (d) logarithmic scales, by May 15, 2020. The day *n* indicates the number of days from December 31, 2019, so that day 1 indicates January 1, 2020.

**Figure A.2:**
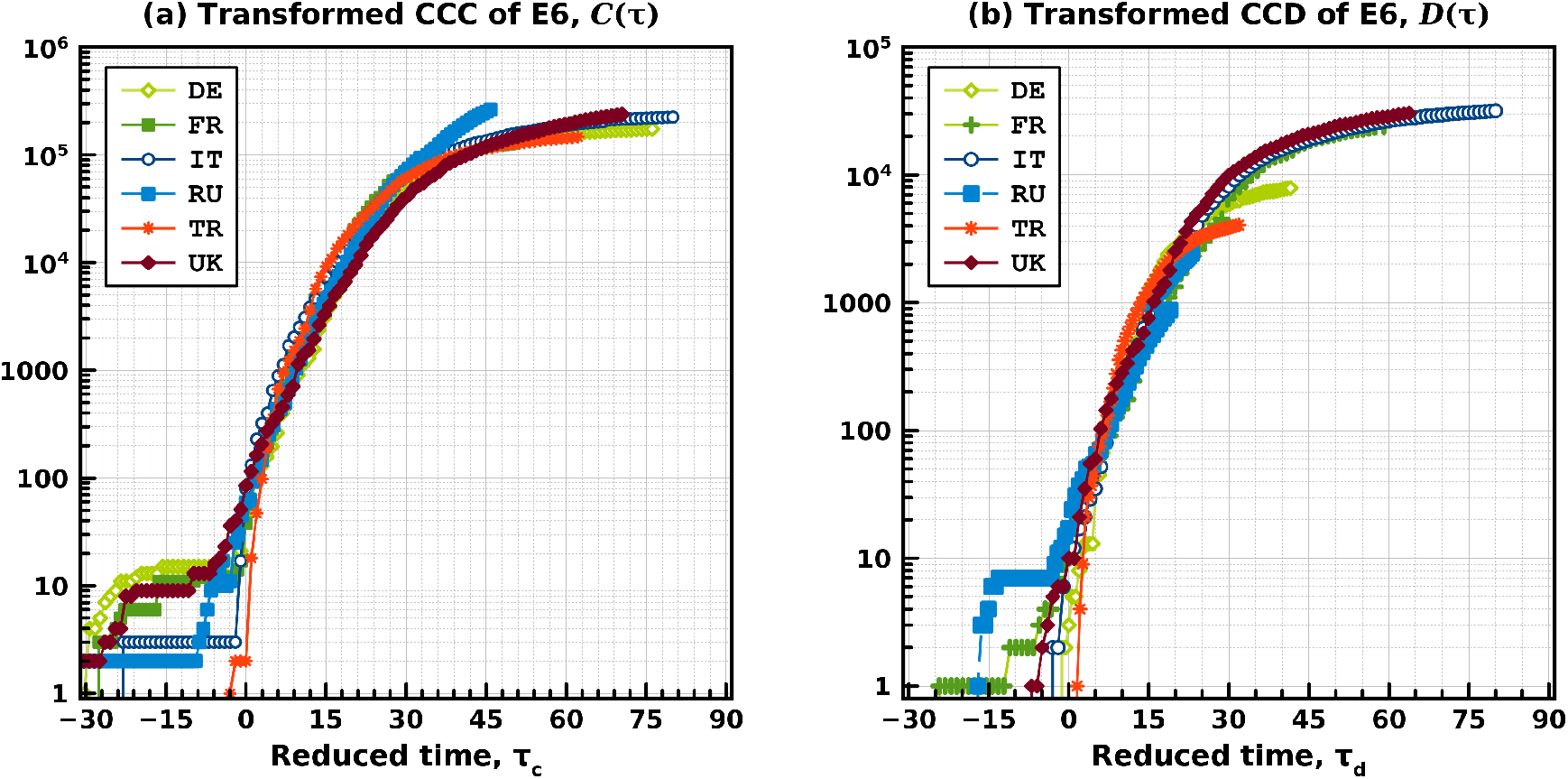
Transformed (a) CCC and (b) CCD profiles of the six nations (E6) in the European continent, by May 15, 2020. The raw data of CCC and CCD are identical to those used on Fig. 1. The reduced time *τ*_*c*_ and *τ*_*d*_ are calculated with Eqs. (4) and (5), respectively, with specific parameter values of *m* and *β* calculated in this study. (See Table 1 for details.) The double arrow toward the end-point of Italy’s profile in (a) indicates the estimated time distance for the U.K. to reach the same CCC level as that of Italy.

## Reference

1 World Health Organization (WHO). Novel Coronavirus – China, Disease outbreak news: 12 January 2020; 2020. URL: https://www.who.int/csr/don/12-january-2020-novel-coronavirus-china/en/.

2 Leung K, Wu JT, Liu D, Leung GM. First-Wave Covid-19 Transmissibility and Severity in China Outside Hubei After Control Measures, and Second-Wave Scenario Planning: a Modelling Impact Assessment. The Lancet. 2020;395(10233):1382–1393. URL: https://doi.org/10.1016/S0140-6736(20)30746-7.

3 Cowling BJ, Ali ST, Ng TWY, Tsang TK, Li JCM, Fong MW, et al. Impact assessment of non-pharmaceutical interventions against coronavirus disease 2019 and influenza in Hong Kong: an observational study. The Lancet Public Health. 2020;5(5):E279–E288. URL: https://doi.org/10.1016/s2468-2667(20)30090-6.

4 Carcione JM, Santos JE, Bagaini C, Ba J. A simulation of a COVID-19 epidemic based on a deterministic SEIR model. arXiv; 2020. URL: https://arxiv.org/abs/2004.03575.

5 Chang SL, Harding N, Zachreson C, Cliff OM, Prokopenko M. Modelling transmission and control of the COVID-19 pandemic in Australia. arXiv; 2020. URL: https://arxiv.org/abs/2003.10218.

6 COVID-19 National Incident Room Surveillance Team. COVID-19, Australia: Epidemiology Report 6: Reporting Week Ending 1900 AEDT 7 March 2020. Communicable Diseases Intelligence. 2020 mar;44. URL: https://doi.org/10.33321/cdi.2020.44.21.

7 The Lancet. COVID-19 in Brazil: “So what?”. The Lancet. 2020 may;395(10235):1461. URL: https://doi.org/10.1016/s0140-6736(20)31095-3.

8 Marchand-Senécal X, Kozak R, Mubareka S, Salt N, Gubbay JB, Eshaghi A, et al. Diagnosis and Management of First Case of COVID-19 in Canada: Lessons applied from SARS. Clinical Infectious Diseases. 2020 03;Ciaa227. URL: https://doi.org/10.1093/cid/ciaa227.

9 Khan N, Fahad S. Critical Review of the Present Situation of Corona Virus in China. SSRN Electronic Journal. 2020; URL: https://doi.org/10.2139/ssrn.3543177.

10 Khan N, Faisal S. Epidemiology of Corona Virus in the World and Its Effects on the China Economy. SSRN Electronic Journal. 2020; URL: https://doi.org/10.2139/ssrn.3548292.

11 The Lancet. Sustaining Containment of COVID-19 in China. The Lancet. 2020 apr;395(10232):1230. URL: https://doi.org/10.1016/s0140-6736(20)30864-3.

12 Stafford N. Covid-19: Why Germany’s case fatality rate seems so low. BMJ. 2020;369. URL: https://www.bmj.com/content/369/bmj.m1395.

13 Fontanet A, Tondeur L, Madec Y, Grant R, Besombes C, Jolly N, et al. Cluster of COVID-19 in Northern France: a Retrospective Closed Cohort Study. MedRxiv. 2020 apr; URL: https://doi.org/10.1101/2020.04.18.20071134.

14 Djalante R, Lassa J, Setiamarga D, Sudjatma A, Indrawan M, Haryanto B, et al. Review and Analysis of Current Responses To COVID-19 in Indonesia: Period of January To March 2020. Progress in Disaster Science. 2020 apr;6:100091. URL: https://doi.org/10.1016/j.pdisas.2020.100091.

15 Lancet T. India Under COVID-19 Lockdown. The Lancet. 2020 apr;395(10233):1315. URL: https://doi.org/10.1016/s0140-6736(20)30938-7.

16 Remuzzi A, Remuzzi G. COVID-19 and Italy: What Next? The Lancet. 2020 apr;395(10231):1225–1228. URL: https://doi.org/10.1016/s0140-6736(20)30627-9.

17 Fanelli D, Piazza F. Analysis and Forecast of COVID-19 Spreading in China, Italy and France. Chaos, Solitons & Fractals. 2020 may;134:109761. URL: https://doi.org/10.1016/j.chaos.2020.109761.

18 Porcheddu R, Serra C, Kelvin D, Kelvin N, Rubino S. Similarity in Case Fatality Rates (CFR) of COVID-19/SARS-COV-2 in Italy and China. The Journal of Infection in Developing Countries. 2020 feb;14(02):125–128. URL: https://doi.org/10.3855/jidc.12600.

19 Mizumoto K, Kagaya K, Zarebski A, Chowell G. Estimating the Asymptomatic Proportion of Coronavirus Disease 2019 (COVID-19) Cases on Board the Diamond Princess Cruise Ship, Yokohama, Japan, 2020. Eurosurveillance. 2020 mar;25(10). URL: https://doi.org/10.2807/1560-7917.es.2020.25.10.2000180.

20 Shim E, Tariq A, Choi W, Lee Y, Chowell G. Transmission Potential and Severity of COVID-19 in South Korea. International Journal of Infectious Diseases. 2020 apr;93:339–344. URL: https://doi.org/10.1016/j.ijid.2020.03.031.

21 Vivanco-Lira A. Predicting COVID-19 distribution in Mexico through a discrete and time-dependent Markov chain and an SIR-like model. arXiv; 2020. URL: https://arxiv.org/abs/2003.06758.

22 de León U, Pérez Á, Avila-Vales E. A data driven analysis and forecast of an SEIARD epidemic model for COVID-19 in Mexico. arXiv; 2020. URL: https://arxiv.org/abs/2004.08288.

23 Sukhankin S. Covid-19 As a Tool of Information Confrontation: Russia’s Approach. SSRN Electronic Journal. 2020 Apr;13. URL: https://ssrn.com/abstract=3566689.

24 Ebrahim SH, Memish ZA. COVID-19: Preparing for Superspreader Potential Among Umrah Pilgrims To Saudi Arabia. The Lancet. 2020 mar;395(10227):e48. URL: https://doi.org/10.1016/s0140-6736(20)30466-9.

25 Gilbert M, Pullano G, Pinotti F, Valdano E, Poletto C, Boëlle PY, et al. Preparedness and Vulnerability of African Countries Against Importations of COVID-19: a Modelling Study. The Lancet. 2020 mar;395(10227):871–877. URL: https://doi.org/10.1016/s0140-6736(20)30411-6.

26 Nkengasong JN, Mankoula W. Looming Threat of COVID-19 Infection in Africa: Act Collectively, and Fast. The Lancet. 2020 mar;395(10227):841–842. URL: https://doi.org/10.1016/s0140-6736(20)30464-5.

27 Ş ahin M. Impact of Weather on COVID-19 Pandemic in Turkey. Science of The Total Environment. 2020 aug;728:138810. URL: https://doi.org/10.1016/j.scitotenv.2020.138810.

28 Peto J, Alwan NA, Godfrey KM, Burgess RA, Hunter DJ, Riboli E, et al. Universal Weekly Testing As the UK COVID-19 Lockdown Exit Strategy. The Lancet. 2020 may;395(10234):1420–1421. URL: https://doi.org/10.1016/s0140-6736(20)30936-3.

29 The Lancet. COVID-19 in the USA: a Question of Time. The Lancet. 2020 apr;395(10232):1229. URL: https://doi.org/10.1016/s0140-6736(20)30863-1.

30 Lescure FX, Bouadma L, Nguyen D, Parisey M, Wicky PH, Behillil S, et al. Clinical and Virological Data of the First Cases of COVID-19 in Europe: a Case Series. The Lancet Infectious Diseases. 2020 mar;20. URL: https://doi.org/10.1016/s1473-3099(20)30200-0.

31 Chinazzi M, Davis JT, Ajelli M, Gioannini C, Litvinova M, Merler S, et al. The Effect of Travel Restrictions on the Spread of the 2019 Novel Coronavirus (COVID-19) Outbreak. Science. 2020 mar;p. eaba9757. URL: https://doi.org/10.1126/science.aba9757.

32 Du Z, Wang L, Cauchemez S, Xu X, Wang X, Cowling BJ, et al. Risk for Transportation of Coronavirus Disease From Wuhan To Other Cities in China. Emerging Infectious Diseases. 2020 may;26(5):1049–1052. URL: https://doi.org/10.3201/eid2605.200146.

33 Aviv-Sharon E, Aharoni A. Forecasting COVID-19 Pandemic Severity in Asia. medRxiv. 2020 may; URL: https://doi.org/10.1101/2020.05.15.20102640.

34 Kinross P, Suetens C, Dias JG, Alexakis L, Wijermans A, Colzani E, et al. Rapidly Increasing Cumulative Incidence of Coronavirus Disease (COVID-19) in the European Union/european Economic Area and the United Kingdom, 1 January To 15 March 2020. Eurosurveillance. 2020 mar;25(11). URL: https://doi.org/10.2807/1560-7917.es.2020.25.11.2000285.

35 Jit M, Jombart T, Nightingale ES, Endo A, Abbott S, and WJE. Estimating Number of Cases and Spread of Coronavirus Disease (COVID-19) Using Critical Care Admissions, United Kingdom, February To March 2020. Eurosurveillance. 2020 may;25(18). URL: https://doi.org/10.2807/1560-7917.es.2020.25.18.2000632.

36 Liu Z, pierre magal, Seydi O, Webb G. Predicting the Cumulative Number of Cases for the COVID-19 Epidemic in China From Early Data. arXiv. 2020 mar; URL: https://doi.org/10.1101/2020.03.11.20034314.

37 Manchein C, Brugnago EL, da Silva RM, Mendes CFO, Beims MW. Strong Correlations Between Power-Law Growth of COVID-19 in Four Continents and the Inefficiency of Soft Quarantine Strategies. Chaos: An Inter-disciplinary Journal of Nonlinear Science. 2020 apr;30(4):041102. URL: https://doi.org/10.1063/5.0009454.

38 Wang X, Tian W, Lv X, Shi Y, Zhou X, Yu W, et al. Effects of Chinese Strategies for Controlling the Diffusion and Deterioration of Novel Coronavirus-Infected Pneumonia in China. medRxiv. 2020 mar; URL: https://doi.org/10.1101/2020.03.10.20032755.

39 European Centre for Disease Prevention and Control. Download today’s data on the geographic distribution of COVID-19 cases worldwide;. Accessed: 2020-05-25. https://www.ecdc.europa.eu/en/publications-data/download-todays-data-geographic-distribution-covid-19-cases-worldwide.

40 GNU P. Free Software Foundation. BASH (Born Again Shell) (3.2. 48) [Unix shell program]; 2016. URL: https://www.gnu.org/software/bash/.

41 Fenlason J, Lord T, Pizzini K, Bonzini P. sed (stream editor); 2017. URL: https://www.gnu.org/software/sed/.

42 Aho A, Weinberger P, Kernighan B. AWK; 2017. URL: https://www.gnu.org/software/sed/.

43 Eaton JW, Bateman D, Hauberg S, Wehbring R. GNU Octave version 4.2.1 manual: a high-level interactive language for numerical computations; 2017. URL: https://www.gnu.org/software/octave/doc/v4.2.1/.

44 The Document Foundation, LibreOffice [Free Office Suite]; 2020. URL: https://www.libreoffice.org/.

45 Kim AS. A Two-Interface Transport Model With Pore-Size Distribution for Predicting the Performance of Direct Contact Membrane Distillation (DCMD). Journal of Membrane Science. 2013;428:410–424. URL: https://doi.org/10.1016/j.memsci.2012.10.054.

